# Resource Overlap and Reporting of Independence in Public Prostate Cancer Research

**DOI:** 10.64898/2026.09.10.26362786

**Authors:** Wenchang Yue, Rathhan Pemma, Ashutosh K. Tewari, Babu J. Padanilam

## Abstract

**Importance:** Public prostate cancer data may appear under different repository identifiers or releases while representing overlapping patients, specimens, or sample records. The implications for reported analytic independence require assessment at both resource and publication levels.

**Objective:** To characterize supported nonindependence among selected public prostate cancer resources and assess reported overlap control in publications using related resources.

**Design:** Cross-sectional methodological audit. Resource records were reconciled through September 2, 2026; recovery of retained sources and adjudication continued through September 8, 2026. Language-model extraction and targeted original-source checks informed deterministic classification. The authors manually verified all positive findings, including supplementary materials, in September 2026.

**Setting:** Public prostate cancer repositories and Europe PMC and PubMed retrieval, with a publication cutoff of August 15, 2026.

**Participants:** A nonrepresentative frame of 189 repository study records and 108 publication/version records contributing 230 publication–resource-pair records.

**Exposures:** Analytic use of both endpoints of a confirmed resource relation in the same linked claim.

**Main Outcomes and Measures:** Supported identity levels and resolvable patient-identifier counts; reported overlap control when linked analyses required independent records. Unresolved evidence was retained separately. Published results were not reanalyzed.

**Results:** The audit identified 101 supported resource pairs: 97 with direct patient, specimen, or sample-record identity, 1 with component containment, and 3 with study/source nonindependence. Among 108 publication/version records, 8 (6 journal articles and 2 preprint versions) used related resources in analyses requiring independence without reporting applicable overlap controls. Another 3 reported applicable controls, 16 retained reporting gaps, and 81 fell outside the qualifying use or relation criteria. One of the 8 had 45 shared sample identifiers in two validation sheets, corroborating sample-record reuse but not donor identity. No global independent-patient total or prevalence estimate was established.

**Conclusions and Relevance:** Resource nonindependence coexisted with absent claim-specific control reporting, with direct sample-record corroboration in one publication. The findings support provenance-aware use and explicit reporting, without establishing effects on published results.

## Introduction

Public prostate cancer datasets support secondary research across disease stages and molecular platforms.^1–3^ Their identifiers describe repository objects, not a uniform biological unit. Projects, cases, specimens, assays, and derived representations have different meanings across repositories.^4–8^ Overlapping portal studies can therefore lead to repeated counting of samples.^9^ A different study name or release label does not establish independent patients.

Hidden transcriptome duplicates and shared-sample networks have been documented previously.^10–12^ Expression similarity can identify possible duplicates but cannot establish donor identity by itself.^13^ Genotype-based methods provide stronger identity evidence across assays and can retain inconclusive matches.^14–18^ Incomplete public metadata further complicate reconstruction of biological sources.^19^

Nonindependence matters when a particular claim requires independent observations. External validation should involve new patients, and evidence synthesis should account for overlapping populations or dependent effects.^20–24^ Repeated records from one individual require appropriate grouping in prediction analyses.^25–27^ However, related resources may legitimately support matched multi-assay analyses or make only one combined contribution. Joint appearance of two resource names is insufficient to determine methodological compatibility.

Prostate cancer resources include harmonized single-cell collections and large primary and advanced-disease cohorts.^28–31^ Related work has also examined cross-resource contamination in pathology benchmarks.^32^ Prostate cancer is informative because a few heavily reused cohorts connect research across disease stages and platforms. Adjudicating each record required disease-area expertise to distinguish assay relationships and clinical context, including whether records represented the same patients or different biological units. We audited documented resource relations, biological counting units, downstream linked use, and reported overlap control. The objective was to assess reported methods and provide source-specific use guidance, not to estimate effects on published results or develop a general duplicate detector.

## Methods

### Design and resource frame

We conducted a resource-first audit of public prostate cancer bulk transcriptomics, identity-informative bulk genomics, and single-cell RNA sequencing. The resource frame was reconciled through September 2, 2026. The mixed design combined complete prostate-specific screens of 31 cBioPortal and 13 GDC records with the earliest accession-ordered records from GEO, BioProject, and dbGaP. These comprised the first 100 GEO screening records plus 4 controls, the first 25 BioProject records plus 3 controls, and the first 10 dbGaP records plus 3 controls. Of 104 GEO records, 101 had accession numbers below 50000 (median numeric accession, 5953). Linked SRA, BioSample, and publication records informed provenance. This selected frame was designed to examine different representations and known relations, not estimate repository-wide prevalence. Eligibility required qualifying patient-derived prostate cancer material; ineligible and unresolved records were retained separately (eTables 1 and 2).

We distinguished patient, specimen, lesion/site, time point, aliquot/library, assay, sample accession, study accession, and publication (Figure 1; eTable 3). Formal positive relations required official metadata, identifier crosswalks, explicit source publications or supplements, or identity-informative molecular evidence. Expression similarity alone was insufficient. A fixed rule assigned direct patient/specimen/sample-record identity, component containment without member identity, or study/source nonindependence. Complete official GEO Sample membership established sample-record identity, not patient counts (eTables 4 and 5).

**Figure 1.**
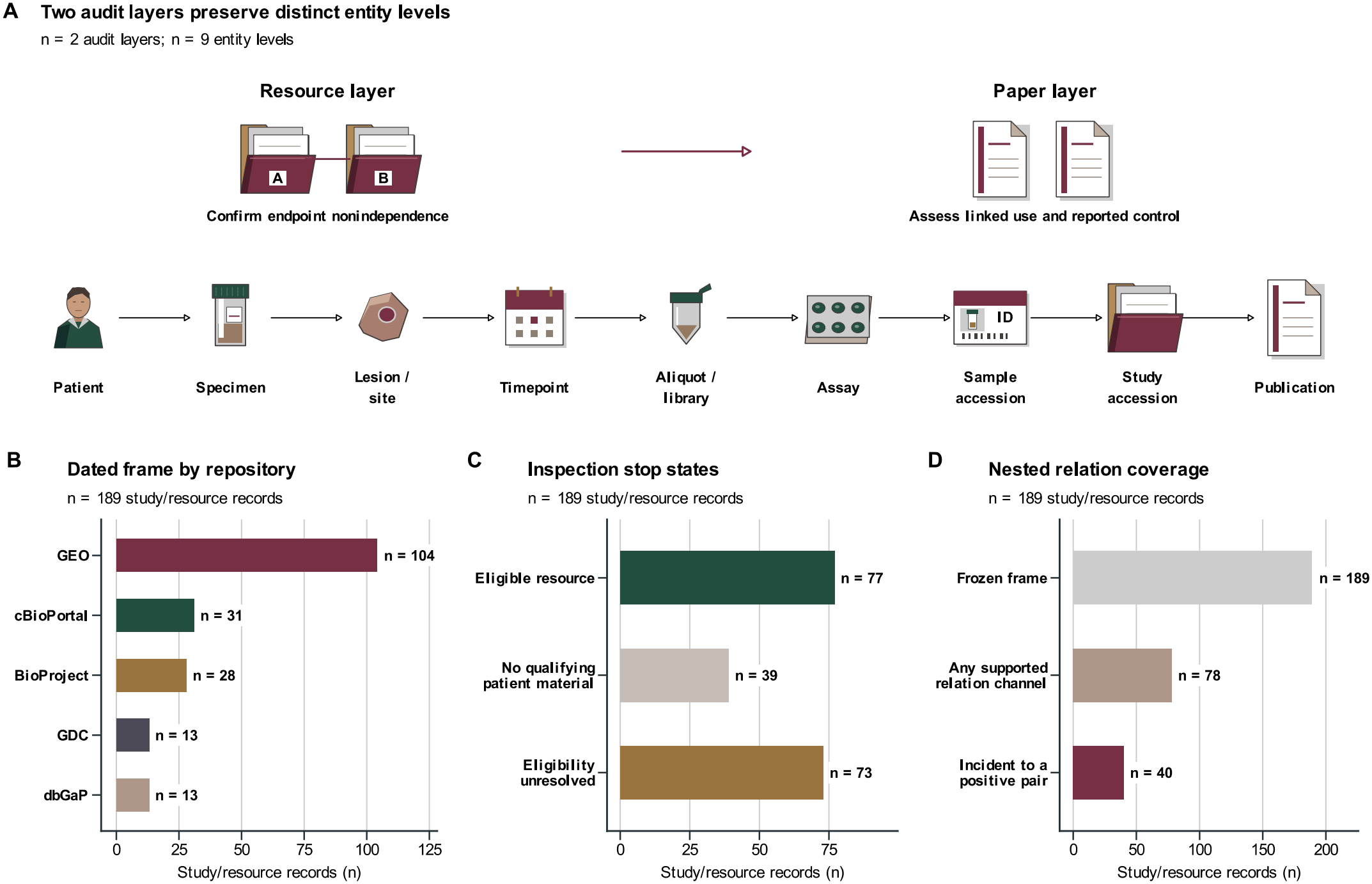
Study design preserves biological identity levels within a dated prostate-cancer resource frame. (A) Resource-relation confirmation precedes paper assessment (n = 2 audit layers), preserving the n = 9 biological and repository identity levels shown. (B) Repository composition of the selected frame (n = 189 study/resource records). (C) Eligibility of those records. (D) Nested coverage: the full frame, records with a supported relation-evidence channel (n = 78), and records incident to a confirmed pair (n = 40). Bars count versioned repository study/accession records, not patients or independent cohorts. An available evidence channel does not itself establish overlap. Counts describe this nonrepresentative frame, not repository-wide prevalence. GEO, Gene Expression Omnibus; GDC, Genomic Data Commons; dbGaP, Database of Genotypes and Phenotypes.

Patient-identifier resolution was examined in every connected resource group. Whole-group unions required coverage of all included endpoints within the stated identifier namespace. Otherwise, counts remained named-subset unions, aggregate declarations, conflicting identities, or study/assay-limited quantities. Pair and subset counts were not added to estimate independent patients globally (eTable 6).

### Publication retrieval and linked-use assessment

Exact identifiers and source-bound aliases were queried through Europe PMC and PubMed with a publication cutoff of August 15, 2026. Records were joined by PMID, PMCID, normalized DOI or provider identifier. Mapping both endpoints of a confirmed relation defined the localized set. Recovered pairs underwent claim assessment. Analytic scope distinguished original prostate-cancer analyses/resources, reviews and pan-cancer work after non-direct relation exclusions. Inspected preprint versions were retained with explicit journal links, without substituting unassessed journal bodies. Scope and procedural classifications remained separate. This was not a study-family census or prevalence denominator; informal names, citation-only references and unexamined materials could be missed (eTable 7).

The bounded evidence set included the main article, Methods, Results, Data Availability, claim-relevant supplements, and linked public code capable of documenting selection or overlap handling. Missing decision-relevant material, retrieval failure, or ambiguous analytic contribution remained unassessable. Matrices or images incapable of containing relevant control statements did not make the set incomplete. After checking these conventional sources, absent statements were recorded as not reported; they did not trigger indefinite searching. Unclear analytic allocation or safeguard applicability was recorded as a reporting gap, separately from unavailable material.

Four sequential questions addressed dual analytic contribution, a shared linked claim, its independence requirement, and evidence of an applicable safeguard. Qualifying safeguards included documented nonoverlapping selection, exclusion, deduplication, patient grouping, or an equivalent claim-specific procedure. A published selection roster could establish exact-record exclusion even without an explicit explanation of overlap. Separate categories retained no dual analytic use, no independence requirement, and explicitly documented single contributions or constituent representations (eTables 8 and 9).

*Procedural misuse as reported* was defined as an established independence-requiring linked use with no qualifying safeguard found in complete bounded public materials. The designation concerns reporting, not intent, misconduct, retention of duplicated records, or result validity. An applicable control statement or original selection roster could establish control without requiring unrelated materials; uncertain contribution or control remained unassessable rather than positive.

### Extraction, adjudication, and descriptive analysis

Evidence excerpts linked to file hashes were initially read using gpt-5.6-sol at high reasoning effort through codex-cli 0.153.0. Fourteen fact fields required source-block locators. Resource tiers came from retained source evidence, and a deterministic rule assigned pair and publication classifications. Targeted follow-up checked original Methods, Results, supplementary captions, tables and published sample selections. Corrections distinguished integrated gene lists, constituent resources, portal representations and independent record contributions. A scoped read-only Claude review informed source adjudication; model agreement was not an acceptance criterion. Versioned prompts, software, source locators and correction history are retained in the eMethods and eTables 7, 10, 11 and 12.

The counting unit was the canonical publication/version record, not an independently reconciled study family. Any qualifying no-control pair determined that record’s classification; otherwise unresolved direct-identity evidence took precedence over reported control, followed by outside categories. A record could contribute several pair rows but only one publication-level classification. Display categories were directly corroborated duplicated inclusion, applicable reported control, and unreported or insufficient handling. The last comprised qualifying no-control findings without direct corroboration and unresolved reporting gaps. Records outside qualifying use or direct identity were excluded from this display.

We reported counts and reasons for unresolved classification. No prevalence fraction or confidence interval was calculated because retrieval coverage and the eligible denominator were not finalized. Original published sample identifiers were compared only to verify selection or reuse, without recalculating outcomes. No hypothesis testing, stochastic sampling, model fitting or outcome reanalysis was performed. Seed 42 was specified for any stochastic step. In September 2026, the authors manually reviewed all 8 positive publication/version records, including supplementary materials, and confirmed the reported-method classifications. This targeted verification did not estimate extraction accuracy across the full set.

### Intended use, reproducibility, and ethics

Use rules linked each relation to the claim, supported identity unit, affected identifiers or grouping key, required action, and versioned source evidence. Acceptance with controls required verified applicability; unresolved evidence required review; rejection applied to the specified independent use, not every use of the resource. Absence of a registry match alone did not establish acceptance (eTables 13 and 14).

The study used public metadata and publications, recruited no participants, and performed no clinical intervention. Ethics approval was not required. Source tables, code, and SHA-256 manifests are retained locally; persistent public deposition remains pending.

## Results

### Resource records and biological units

Before assessing paper use, we examined the biological identity levels represented in the resource frame (Figure 1A), which comprised 189 study records: 104 GEO, 28 BioProject, 31 cBioPortal, 13 GDC, and 13 dbGaP records (Figure 1B). Seventy-seven records were eligible, whereas 39 lacked qualifying patient-derived material and 73 remained unresolved (Figure 1C; eFigure 1A). Within the full set of 189 records, 78 had supported relation-evidence channels; of these, 40 were incident to a confirmed pair (Figure 1D; eFigure 1B; eTable 15).

### Supported cross-resource relations

The 101 supported pairs connected 62 dataset identifiers in 15 groups (Figure 2A; eFigure 2). To relate this graph to the resource frame, we matched 37 of the 40 related frame records directly and 3 through project/version crosswalks, representing 39 graph identifiers. Frame records and graph nodes were therefore different counting units (eTable 1).

**Figure 2.**
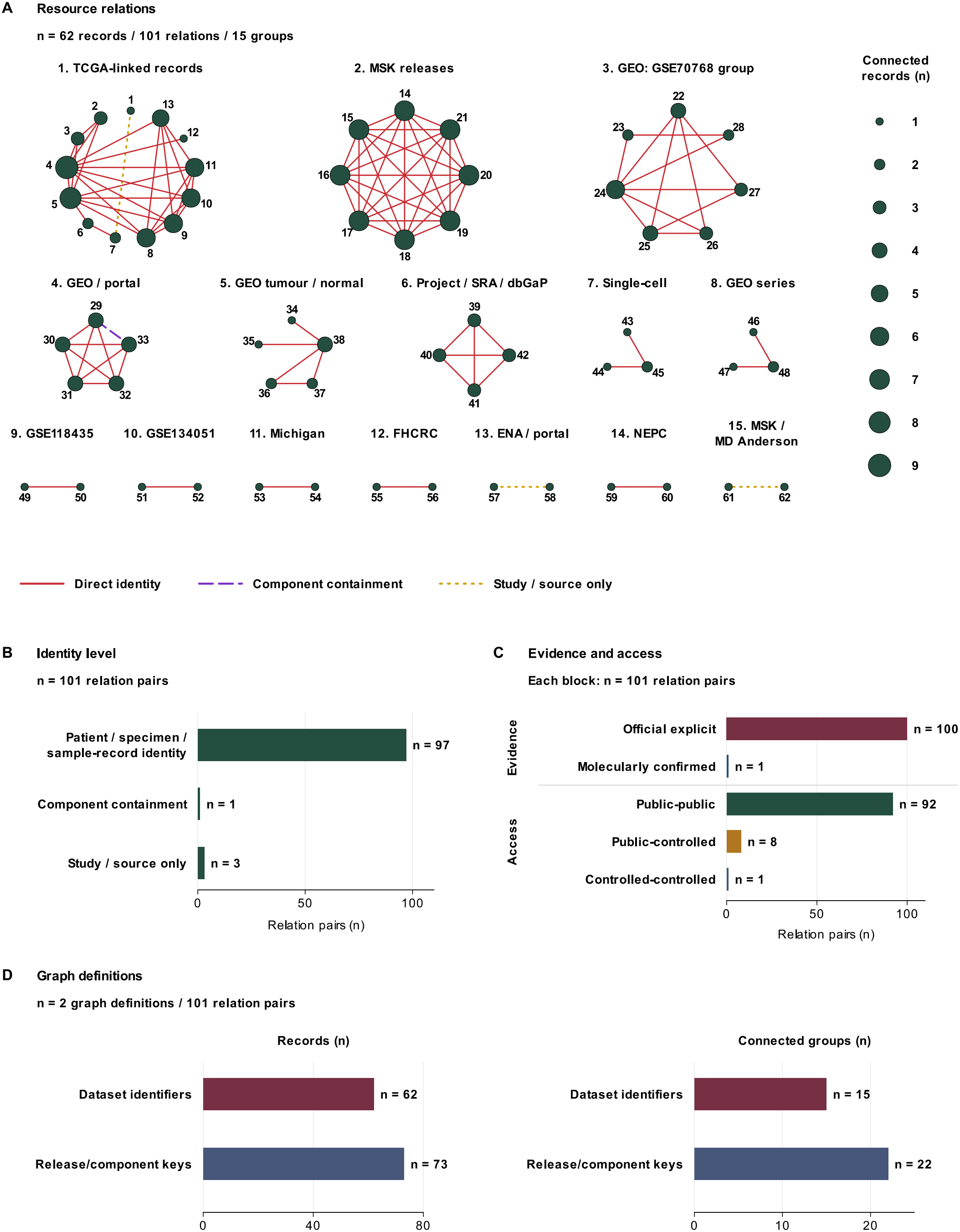
Supported resource relations form connected groups across public data records Legend continued. **(A)** All n = 62 study/project or dataset-release records and n = 101 supported pairs in 15 connected groups. Bubble area represents degree, the number of directly connected records, not patient count. Small record numbers and bold group numbers match the names and platforms in eFigure 2. Solid red lines indicate direct patient, specimen or sample-record identity; dashed purple lines indicate component containment without member-level identity; dotted golden-yellow lines indicate study/source nonindependence. Relations are undirected; layout and distance have no quantitative meaning. Missing edges do not establish independence, and a connected group does not imply all-to-all patient overlap. **(B)** Relation categories for n = 101 pairs. (C) Evidence source and endpoint access status, each classifying the same n = 101 pairs. Official or molecular evidence describes how a relation was established; public or controlled access describes its endpoints. (D) Node and connected-group counts under two definitions: dataset identifier and release/component key. Pair counts are nonadditive and do not estimate independent patients. Exact identifiers, platforms and supporting sources are provided in eFigure 2 and eTable 4.

Ninety-seven pairs supported direct identity: 68 patient-level, 1 specimen-level, and 28 sample-record-level relations. One additional pair supported component containment without member identity, and 3 supported study/source nonindependence (Figure 2B). Official evidence supported 100 pairs and molecular evidence 1; access classes were public–public for 92, public–controlled for 8, and controlled–controlled for 1 (Figure 2C; eTable 16). Keeping release/component distinctions produced 73 nodes in 22 groups over the same 101 pairs (Figure 2D).

### Patient-count resolution across all groups

All 15 groups were assessed for patient-count resolution: three supported whole-group public patient-identifier unions, 7 only named subsets, 2 aggregate declarations, 1 conflicting identity fields, and 2 study/assay-limited evidence (Figure 3A; eTable 6A). At the pair level, 97 quantities were exact at the recorded unit, 2 were lower bounds, and 2 were unknown. The 97 exact-magnitude and 97 direct-identity pairs were, however, different sets, intersecting at 96 pairs.

**Figure 3.**
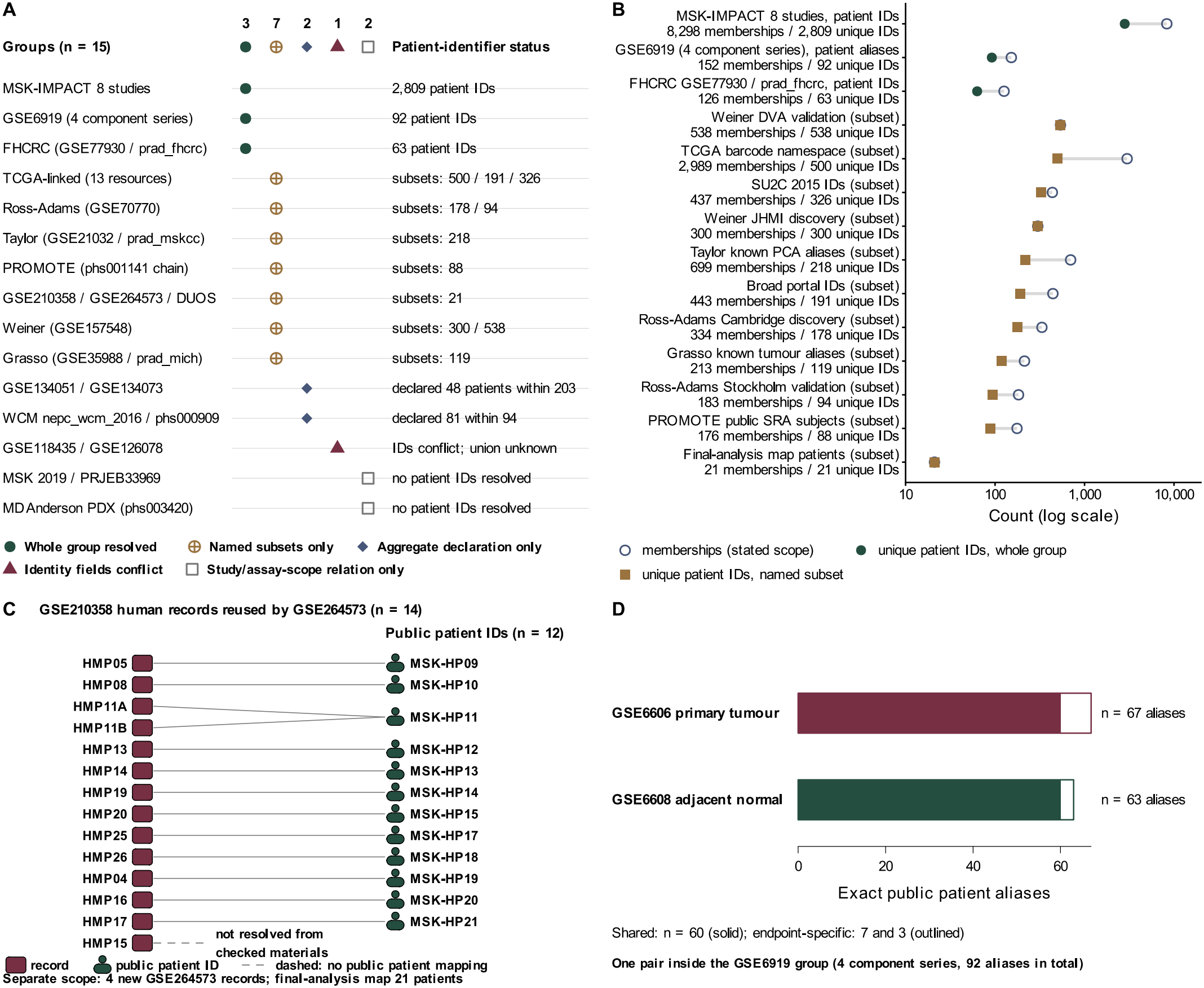
Patient-identifier resolution across supported resource groups. (A) Patient-identifier resolution for all n = 15 connected groups, classified by the scope of available identity evidence. Unknown quantities are stated as text, not zero. (B) n = 14 exact identifier-union rows from 10 groups, comprising 3 whole-group and 11 named-subset rows. On the logarithmic axis, open circles show summed endpoint memberships; filled marks show unique patient identifiers within each stated scope. Equality means no repeated identifier across those memberships. Counts are observed public identifiers, not molecularly confirmed donors, and are nonadditive across rows. (C) Of n = 14 GEO sample records reused from GSE210358 in GSE264573, 13 map to n = 12 patient identifiers; a dashed line marks the unresolved record. The 4 new records and 21-patient final-analysis map have a separate scope and do not resolve that record. (D) One component pair in the GSE6919 group: GSE6606 (n = 67 patient aliases) and GSE6608 (n = 63) share 60 aliases. Solid segments show shared aliases; outlined segments show endpoint-specific aliases. This pair is distinct from the four-series group union. Color identifies categories or endpoints, with shapes or labels providing redundant distinctions. GEO, Gene Expression Omnibus.

Fourteen rows from 10 groups supported exact identifier unions: 3 whole-group and 11 subset rows (Figure 3B). Among these, the eight-resource MSK group had 8298 endpoint patient memberships but 2809 public identifiers. The corresponding totals were 152 memberships and 92 identifiers for the four-series microarray group, and 126 memberships and 63 identifiers for the paired FHCRC representations.

For single-cell reuse, 13 of 14 reused repository records mapped to 12 patient identifiers and 1 remained unresolved (Figure 3C); the separate final-analysis subset with 21 patient identifiers did not resolve that unmatched record. At the pair level, resources with 67 and 63 patient aliases in one microarray pair shared 60 aliases (Figure 3D); this was distinct from the whole-group union of 92. All 38 unit-level rows were retained: 27 patient, 8 assay/sample-record, and 3 specimen rows (eFigure 3; eTable 6B).

## Linked paper use and reported overlap control

Retrieval yielded 1504 canonical retrieval records and 3237 publication–relation candidates. After source recovery added 21 publication/version records and 70 pair rows, the localized set totaled 108 records and 230 pair rows; 101 records contained a direct-identity relation (Figure 4A). This set mapped both endpoints of 65 of the 101 resource pairs (Figure 4B), leaving the other 36 pairs without exact representation.

**Figure 4.**
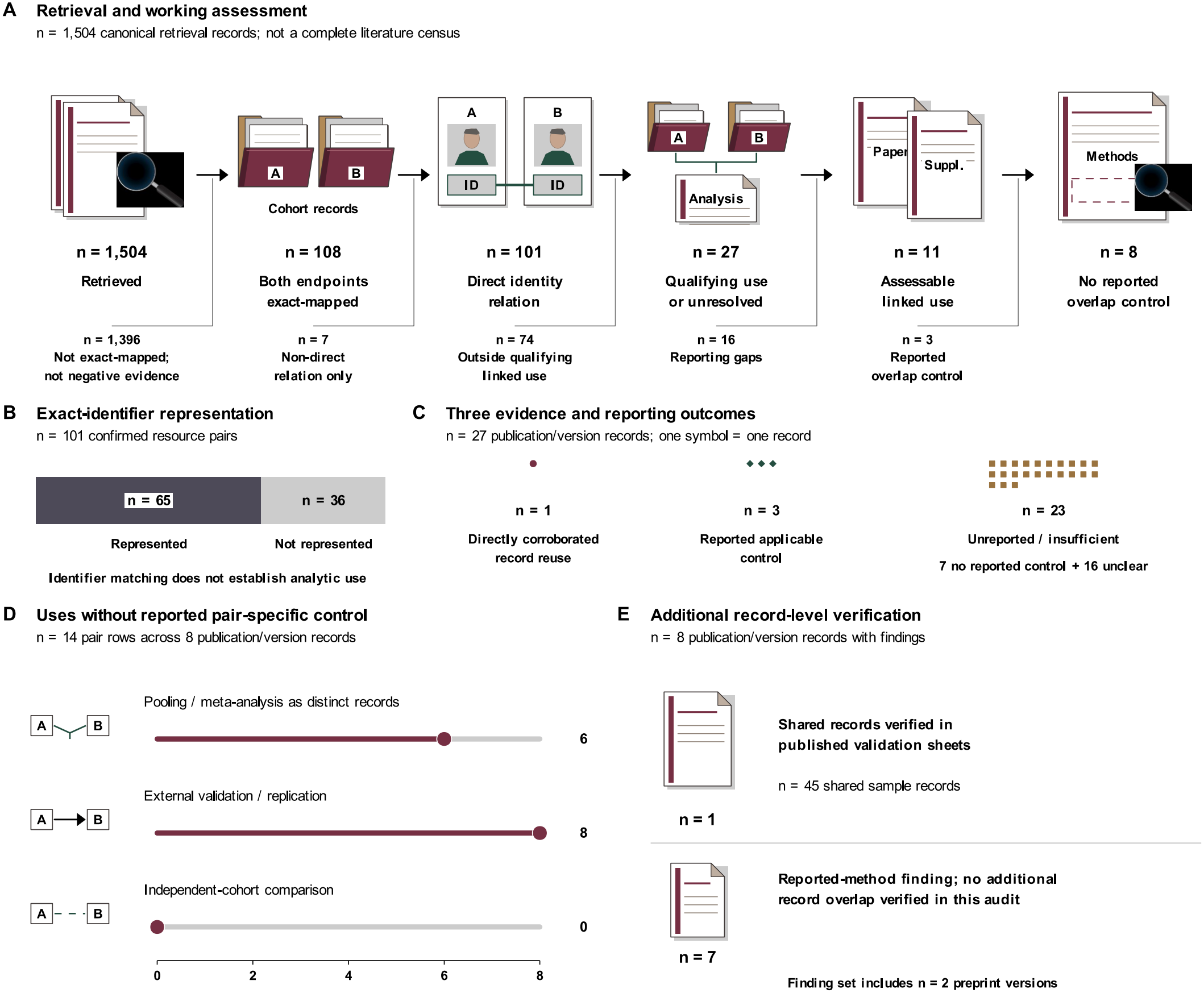
Paper assessment distinguishes reported-method findings from additional record-level evidence Legend continued. (A) Selection and assessment of n = 1504 retrieval records, including n = 108 publication/version records with both resource endpoints mapped. Exits distinguish non-direct relations and uses outside the qualifying linked claim; reporting gaps remain separate from assessable findings. Counts refer to records/versions, not independent study families. Unmapped records are not negative-use findings. (B) Representation of n = 101 supported resource pairs in the localized publication set. Segment lengths show pair counts. Identifier matching does not establish analytic use, and nonrepresentation does not establish absence from the literature. (C) One symbol represents one of n = 27 qualifying or unresolved records. The n = 23 unreported/insufficient category combines 7 no-reported-control findings without direct corroboration and 16 reporting gaps. Together with the n = 1 corroborated record, the 7 findings constitute the strict n = 8; the reporting gaps are not counted as established duplicate use. The n = 3 controlled records may document safeguards in selection rosters without an explicit prose statement. (D) Analysis types among n = 14 no-control pair rows from n = 8 records (6 journal articles and 2 preprint versions). Bar lengths count publication–resource-pair rows, not independent publications. (E) Additional record-level evidence within those n = 8 findings. In one publication, all n = 45 sample identifiers in one validation sheet also occur among n = 64 in another, with matching scores and groups. The other 7 lack this additional corroboration in the audit. Shared sample records do not establish unique-donor counts or training–test overlap. No reported control means none was found in the complete bounded claim-relevant materials; it does not establish author awareness, undisclosed procedures or result invalidity. The authors verified all n = 8 findings, including supplementary materials. No prevalence estimate or outcome reanalysis is presented. Reporting gaps and retrieval coverage are detailed in eFigure 4.

The three-category display comprised 1 record with directly corroborated duplicated inclusion, 3 with applicable reported control, and 23 with unreported or insufficiently described handling (Figure 4C; eTable 17). Of the 23, 7 were qualifying no-control findings without direct corroboration, whereas 16 retained reporting gaps. Another 74 lacked qualifying linked use and 7 had only non-direct relations. Counting the corroborated example with the other qualifying findings, 8 records met the no-reported-control definition: 6 journal records and 2 retained preprints. These findings contributed 14 pair rows, comprising 8 external-validation/replication uses and 6 pooled analyses (Figure 4D).

One journal finding had record-level corroboration: all 45 sample identifiers in one published validation sheet appeared among the 64 in another. This established sample-record reuse between two validations, not 45 unique donors or overlap with their training cohort. For the other 7 findings, however, classification rested on reported methods without direct confirmation of duplicated inclusion (Figure 4E; eTable 18). Among the preprint findings, one reported pooled all-sample contingency analysis and the other reported bulk-cohort validation without applicable overlap safeguards in their bounded public materials.

The prior 31-record review left 15 reporting gaps with complete claim-relevant materials. One added preprint had 3 pairs with unreported allocation and a separate unavailable-caption pair. Thus, 16 records retained reporting gaps: 15 concerned allocation/dependence and 1 the patient-level applicability of a general duplicate-exclusion procedure (eFigure 4A and B).

Of 108 localized records, 96 reported original prostate-cancer analyses or resources (90 journal records and 6 preprint versions; 195 pair rows), 6 were reviews, and 1 was a pan-cancer methods paper without a separate prostate-specific claim. Five records were not assessed for original-analysis scope after exclusion at the non-direct resource-relation step (eFigure 4C).

Of 1504 retrieval records, 1232 had identity-matched, hash-verified raw files: 915 JATS XML, 177 BioC XML, 118 HTML and 22 PDF (eFigure 4D; eTable 7). The remaining 272 were not classified as negative-use findings or unavailable. File identity did not establish reading or supplement coverage. Four newly assessed preprints contributed 45 pairs without adding a no-control or controlled finding. A linked journal article published before cutoff remained unassessed, without substitution or independent counting. Publication-status checks of the preceding 99 records identified no new adverse notices. One scope-compatible journal record with a retraction link remained outside no-control findings.

### Use-specific rules and reusable evidence

These resource-level findings informed restrictions on proposed independent contributions for all 101 supported pairs (Figure 5A). The required action depended on the supported relation: the 97 direct-identity pairs required grouping or exclusion at the supported level. The component relation required selecting one representation or separating its component; the 3 study/source relations required member-level resolution or continued review/restriction (Figure 5B; eTable 14).

**Figure 5.**
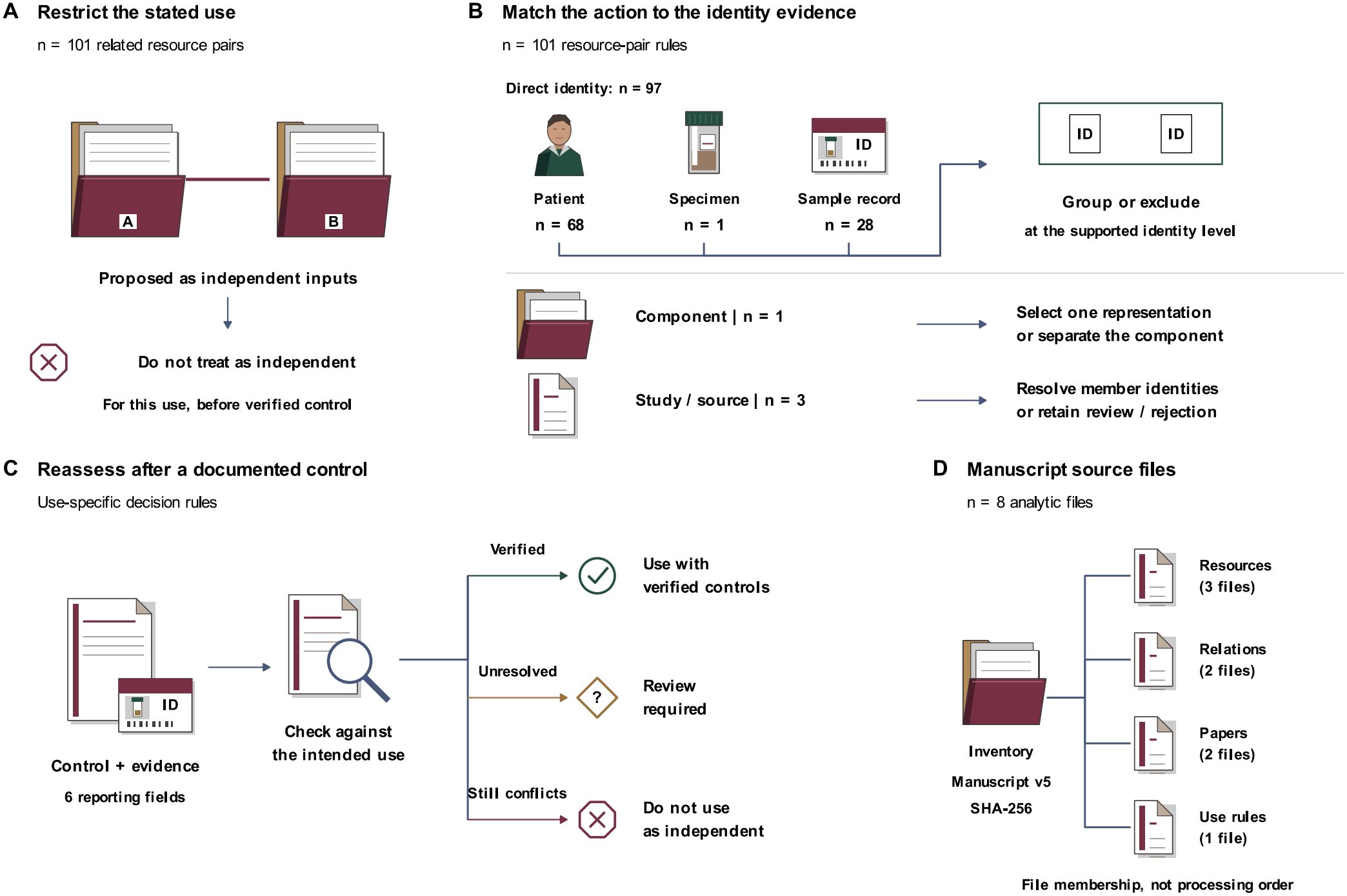
Source-linked rules specify controls for intended independent use. (A) Restrictions for treating n = 101 related resource pairs as independent inputs before applicable controls are verified. These are prospective use rules, not observed paper-method events or blanket rejection of the datasets. Not every pair has identified shared patients. (B) Actions matched to evidence for n = 101 pairs. Direct identity requires grouping or exclusion at the supported patient, specimen or sample-record level. Component containment requires selecting one representation or separating its component; study/source relations require member-level resolution or continued restriction. Lines join parallel action categories, not interchangeable identity levels. Removing duplicate sample records alone does not establish patient independence. (C) Conditional reassessment: verified control supports ACCEPT_WITH_CONTROLS, uncertain applicability requires REVIEW_REQUIRED, and an unresolved independence conflict retains REJECT_FOR_THIS_USE. Shapes distinguish these decisions. They are proposed rules, not observed intervention outcomes. The n = 6 reporting fields are detailed in eFigure 5A; completing fields alone is insufficient. Unrestricted ACCEPT is not assigned in this related-resource workflow. (D) n = 8 analytic files linked to a common SHA-256 inventory. Lines indicate file membership, not processing order. Resource, relation/evidence, paper-audit and use-rule files retain their own counting units. The inventory contains local manuscript sources, not a newly promoted formal release or validated patient-level splits. File contents and counts are listed in eFigure 5B.

Reassessment of a proposed use under these restrictions required 6 fields: exact endpoints/releases; claim/intended use; supported identity level; affected identifiers/grouping key; exclusion/grouping action; and source/version/artifact/hash (Figure 5C; eFigure 5A).

To make these assessments traceable, the local inventory linked 8 analytic files on resources, hierarchy, relations, identity evidence, paper audits and use rules (Figure 5D; eFigure 5B). Within this inventory, the paper tables contained 108 publication/version records and 230 pair rows; each relation file contained 101 pairs.

## Discussion

This audit documented 101 resource relations and examined 108 publication/version records. Eight met the no-reported-control definition: six journal records and two preprints. One was also supported by shared sample identifiers in published validation outputs. We presented that example, 3 applicable controls and 23 unreported or insufficiently described cases separately: the other 7 no-control findings and 16 reporting gaps. It did not establish duplicated inclusion in all 23. These observations demonstrate the phenomenon in the audited material, not its prevalence across prostate cancer research.

Resource reuse is not inherently defective. A cohort may legitimately have several repository representations, releases, or assays. The problem arises when related representations are expected to supply independent evidence without sufficient documentation of how overlap was handled. Subject-level grouping and source-aware evaluation address this distinction.^33^ Incomplete reporting of methodological detail is not confined to secondary data use; among the most cited recent clinical trials, stated intentions to share data and code exceeded what was actually available.^34^ The observed patient-identifier unions illustrate why summing study membership can overcount people, while neither public aliases nor duplicate-file removal establish a global donor union.

Reporting was the endpoint because readers need an account of sample allocation and overlap handling to assess a claim requiring independent data. A resource relation alone cannot answer that question. Reported selection and control procedures allow subsequent investigators to check the relevant identity level before analysis. Authors can also correct incomplete descriptions. Estimating effects on published results would require re-executing analyses, which was outside this audit.

Original-source checks changed several initially positive classifications and resolved previously unclear uses. Combining gene lists from several resources was not equivalent to pooling their patients. Parent and component identifiers could describe one multi-assay contribution, and portal-level replication wording could remain ambiguous. Published selection tables also showed that overlapping resources need not contribute both copies of registered overlapping records. Conversely, original supplementary displays established pooled testing and separate validation that text extraction alone had not resolved. A general duplicate-exclusion statement could still leave its patient-level application across assays unclear.

Model-assisted extraction requires transparent reporting and oversight.^35,36^ Evidence-synthesis studies have reported variable extraction performance and omissions.^37^ Source locators, explicit rules and correction histories make decisions inspectable, but do not establish reading accuracy. Author verification covered all positive findings, but did not measure missed findings or establish independent dual extraction. Agreement between models and reapplication of the classification rules are different checks, and absence of a reported safeguard cannot prove absence of an undisclosed procedure. Findings should therefore remain tied to the inspected source version and claim, with direct sample-record corroboration distinguished from a reporting-based judgment.

Evidence records retain each resource version and its intended use. A useful record identifies the exact representation, biological level, intended use, affected identifiers, and applicable remedy. Persistent data citations and verifiable reusable objects provide an established basis for this approach.^38–40^

## Limitations

The resource frame was dated and nonrepresentative. Known related resources may be overrepresented; missed reports may counter this. Accession ordering may preferentially capture older or platform-split GEO representations. The magnitude of selection bias was not quantified. Recovery of additional exact matches showed that the original mapping was incomplete. Informal names, source-paper citations and unexamined or inaccessible materials could leave further publications undetected. The 272 records without verified raw articles, incomplete supplement/image coverage and bounded rather than exhaustive version-family reconciliation precluded a finalized eligible denominator. A newly assessed preprint had an explicitly linked journal article published before the cutoff that was not independently assessed. Reporting gaps may differ systematically from assessable cases, and targeted corrections do not quantify extraction error across the full set. Two no-control findings concern preprints rather than peer-reviewed reports. Public identifiers may change across platforms, so unmatched names cannot exclude hidden reuse. We did not perform a systematic molecular-fingerprint search or establish a global independent-patient count. Repository documentation can change after the recorded check date. The endpoint was reported methodological compatibility; effects on published results were not measured.

## Conclusions

Documented nonindependence among public prostate cancer resources coexisted with absent applicable control reporting and unclear descriptions of analytic use. One published validation example also contained directly shared sample records. Resource provenance and claim-specific independence checks should precede independent use. No effects on published results were established.

## Data Availability

The supplementary workbook provides the derived resource tables and publication-level audit summaries. The authors permit public sharing of the derived audit data and analysis code. The complete versioned files and SHA-256 manifests are retained locally pending public repository deposition. Copyrighted full texts and restricted individual-level data are excluded from public sharing.

## Acknowledgment of AI Assistance

AI tools, including Claude Opus 5 (Anthropic; xh) and GPT-6 Astra (OpenAI; high), were used through September 2026 to assist with study execution, programmatic checks of the target publications, code review and checks of related materials, and manuscript review and language editing. The authors reviewed the AI-assisted work and take responsibility for the accuracy and integrity of the manuscript.

Author contributions: Wenchang Yue conceived and designed the study, developed the methodology, conducted the investigation and analyses, and wrote the original draft. Rathhan Pemma helped introduce the project and organize the study data. Babu J. Padanilam supervised the study and reviewed and revised the manuscript. Ashutosh K. Tewari provided guidance and secured funding.

Funding/Support: This work was supported by the Seed Grant (0285-5822) from the Department of Urology, Icahn School of Medicine at Mount Sinai.

Role of the Funder/Sponsor: The funder had no role in the study design, data collection and analysis, interpretation of data, writing of the report, or the decision to submit the article for publication.

Conflict of interest disclosures: The authors declare no relevant conflicts of interest.

