## Supplementary methods and figures for "Resource Overlap and Reporting of Independence in Public Prostate Cancer Research"

**Contents**

eMethods; eFigures 1–5; eTables 1–18. Complete tables are supplied in SupplementaryTables.xlsx. The table notes here do not replace the full multi-sheet workbook.

eFigure 1. Repository-specific eligibility and relation evidence within the resource frame
eFigure 2. Full identifiers and hosting platforms of connected public data records
eFigure 3. Unit-specific quantities and unresolved counts across all resource groups
eFigure 4. Material availability, reporting gaps, publication scope and retained-source coverage
eFigure 5. Reporting fields and local manuscript-source files support use-specific reassessment
eTable 1. Frozen public prostate-cancer resource frame
eTable 2. Cohort registry and verified source metadata
eTable 3. Patient-to-publication entity crosswalk
eTable 4. Confirmed resource nonindependence relations
eTable 5. Identity evidence and intended-use fields
eTable 6. Patient-identifier resolution and unit-specific quantities across all resource groups
eTable 7. Retrieval composition, source coverage and classification versions
eTable 8. Paper-level reported overlap-control classifications
eTable 9. Publication-relation evidence ledger
eTable 10. Field-level production provenance for the publication-relation ledger
eTable 11. Correction, review and recovery lineage for the publication-relation ledger
eTable 12. Scope of retained and current bounded checks
eTable 13. Pair-specific independence-preserving use rules
eTable 14. Intended-use decision matrix for confirmed resource relations
eTable 15. Audit frame, counting units and interpretation boundaries
eTable 16. Taxonomy of confirmed resource relations
eTable 17. Descriptive linked-use classifications and analysis types
eTable 18. Additional sample-record evidence for the eight reported-method findings

### Supplementary Methods

The analysis includes 108 publication/version records and 230 pair rows. Historical extraction and review records are retained separately and do not constitute repeated independent validation.

**Resource representation and evidence**

The audit distinguished patients, specimens, lesions/sites, time points, aliquots/libraries, assays, sample accessions, study accessions and publications. Source evidence retained the exact endpoint, version, locator, check date, original artifact and SHA-256. Eligibility, the opportunity to assess a relation and a supported positive relation were separate attributes.

The dated frame comprised n = 189 study records, not independent cohorts. Its 40 frame records involved in supported relations mapped to 39 graph identifiers. The supported registry comprised n = 101 pairs: 97 direct patient/specimen/sample-record relations, 1 component relation without member-level identity, and 3 study/source relations. Official complete GEO Sample membership established sample-record identity rather than patient identity. Exact quantities, lower bounds and unknown magnitudes retained their recorded units. The 97 exact-magnitude and 97 direct-identity sets intersected at 96 pairs.

All 15 dataset-identifier groups underwent patient-resolution assessment. Whole-group unions required coverage of all endpoints in the stated namespace; otherwise quantities remained named subsets, aggregate declarations, conflicts or study/assay-limited counts. The complete tables retain 15 group records and 38 unit-quantity records. Fourteen plotted patient-identifier union rows comprise 3 whole-group and 11 subset rows from 10 groups. They are not a global count of independently verified donors.

**Retrieval and retained-source recovery**

Initial candidate searches were logged on August 3, 2026. Source-bound identifiers and accepted aliases were queried through Europe PMC and PubMed with a publication cutoff of August 15, 2026. Query semantics and provider failures were retained. Records were joined through PMID, PMCID, normalized DOI and provider-local identifiers. The multichannel retrieval frame contained n = 1,504 canonical publications and n = 3,237 publication–relation candidate rows.

The original localized set contained 87 publications/160 pairs. Subsequent retained-source checks examined original articles, structured tables, links and localized identifiers. The final file/identity join verified 1232 of 1504 retrieval records: 915 JATS XML, 177 BioC XML, 118 HTML and 22 PDF files. The preceding BioC/HTML checks covered 634 candidate pair rows and 266 source locations. Later recovery retained 22 additional article bodies and 8 previously retained bodies underwent bounded reading. The remaining 272 records did not have verified raw articles in the joined inventory. They were not negative findings or established as unavailable. File identity did not certify complete reading, supplementary coverage or absence of extraction error (eTable 7).

The first recovery added 10 publications/16 pairs and the next source closure added 2 publications/2 pairs, producing the preserved 99/178 set. The current join adds 9 publication/version keys and 52 pair rows, comprising 7 additional rows with completed source assessment and 45 rows from 4 newly assessed preprint versions. The result is 108 records/230 pairs. Scope counts are 96 original prostate-cancer analyses/resource contributions (90 journal records and 6 retained preprints; 195 pairs), 6 reviews and 1 pan-cancer methods paper (30 pairs combined). Five new records with only non-direct resource relations exit before original-analysis scope assessment (5 pairs); they are explicitly not assessed, not presumed original analyses. Scope and reported-use classifications remain distinct.

**Bounded materials and extraction**

The public evidence set included relevant main text, Methods, Results, Data Availability, supplementary methods/cohort information and linked public code capable of documenting selection or overlap handling. Relevant missing material, failed retrieval or unresolved contributions remained unassessable. Material irrelevant to the claim did not create an automatic exception. All declared supplemental titles and scopes were checked before deciding whether a missing item could affect the claim. Once these conventional sources were covered, an absent statement was recorded as not reported; generic searching was not extended indefinitely. Reporting gaps in analytic allocation, dependence or safeguard applicability were recorded separately from missing source material. Neither absence of a statement nor unclear reporting was interpreted as evidence of author awareness.

Initial extraction used gpt-5.6-sol, high effort, through codex-cli 0.153.0. Fourteen fact fields required source-block locators. Retained prompt SHA-256: 9887eb648d266edf808b3b7530b589f35e5c42862b9b6ba2e1647b48c4bed1f1; schema SHA-256: 6a3768fee39641fabec59649bc7eae6d81c9b182567b6be8384e3e80a548ac8b. Those describe the historical extraction, not a new corpus-wide run. Targeted source checks used original XML/PDF passages, supplementary images, workbook headings/notes and published sample-ID fields. Multiple excerpts from the same passage were not independent sources.

The deterministic rule first excluded non-direct relations, reference-only contributions and unlinked claims. Explicit single contributions or constituent representations and uses not requiring independence remained outside the qualifying category. Qualifying independence-requiring patterns were training/test separation, external validation/replication, independent cohort comparison and pooled analysis as distinct records. A flat source list alone did not prove those contributions.

An affirmative, applicable safeguard or original published selection roster could establish control despite incomplete unrelated materials. A negative safeguard finding required a resolved independence-requiring claim, no applicable constituent-only statement, and complete claim-relevant public materials. Patient grouping, sample-record exclusion and file checksum uniqueness were not interchangeable. Publication-level classification gave precedence to a qualifying no-control pair, then unresolved direct evidence, then reported control, then outside categories. Publications with no direct relation remained separate. One publication/version record could contribute several pair rows but only one publication classification. Exact versions and explicit journal links were preserved; the records are not a fully reconciled census of independent study families.

**Source adjudication and corroboration**

Original-source adjudication corrected several earlier positive classifications. Gene-list consolidation was distinguished from pooled patients, and ambiguous portal replication or source contribution remained unresolved. Published selection data could show that both endpoints of a registered overlap were not selected, even without a generic overlap statement. The source and correction indexes retain these distinctions rather than equating co-mention, resource overlap and duplicated analytic inclusion.

One scoped Claude review examined 14 publications/19 pairs and the code that assigned classifications, returning claude-fable-5-1 at high effort. The main executor adjudicated comments against the originals. This was not a whole-project or 97-publication review. Final preprint adjudication subsequently used the already supplied original raster supplement and all declared material scopes; it was not represented as another reviewer PASS.

A subsequent bounded source review covered all 31 publications that remained unassessable and their 60 associated pair rows, including 53 initially unassessable pairs. Two AI readers examined disjoint case lists and returned source locators; the main executor checked the original evidence and adjudicated the proposals. This was targeted model-assisted source reading, not independent human verification or a repeat whole-corpus extraction. The retained closure table contains 93 evidence locators resolving to 71 cited files. Three publications moved to qualifying use without reported control, 1 to supported exact-record selection control, and 12 outside qualifying use; 15 retained reporting gaps. Twenty-one pair classifications changed across 16 publications.

The newly supported selection control was established from an original roster selecting 14 original records and 4 newly contributed records, without selecting a second copy of the 14 registered reanalysis records. This controlled exact-record reuse for that selection; it did not establish global patient independence or an explicit author explanation of why overlap was excluded. A separate publication reported exclusion of repeated samples, but did not resolve whether the same patients were grouped across assays. Its reporting-gap classification concerns the applicability of a reported control, not a failure to locate a control statement. Another publication documented clustering selection while leaving genomic aggregation unclear; control was assessed by linked claim, not assigned indiscriminately to the whole publication.

For DOI 10.1101/2025.11.16.688689, retained bioRxiv version 1, the main Methods named both MSK releases and described pooled frequencies and Fisher testing. Original supplementary Figure 1A explicitly showed all-sample contingency rows with N = 3,433. Main article and all five supplements were available; no applicable overlap safeguard was found in the bounded claim materials. The finding is limited to that version and reported method. No inference was made that all 259 registered shared public patient IDs were duplicated in its analytic inputs.

For PMID 38441550, published validation output sheets contained 45 and 64 GSM identifiers; all 45 identifiers from the smaller sheet appeared in the larger. This was a comparison of original sample identifiers and reported selection fields, not an outcome reanalysis. It established reuse between the two validation outputs, not 45 verified unique donors or overlap with the training cohort. For the other seven no-control publications, actual duplicated inclusion was not established.

The two most recent recovered records were read from their retained original sources. PMID 29017058 represented one FHCRC/UW cohort at GEO and cBioPortal; its separate independent comparison concerned SU2C/PCF, not the two paired representations. The named pair therefore contributed one cohort rather than two independent cohorts. This did not establish global donor independence or resolve relations with other cohorts. PMID 33043165 was a review whose table cited the two resources in different source-study rows, not an original paired analysis. Neither record added a no-control or controlled finding. Sixteen retained evidence locations include the named-pair Methods, Results, resource-table and supplementary context. No further general retrieval was required for these two decisions.

**Exact-version closure**

The 45 new pair rows received fixed bounded dispositions: 34 descriptive or within-cohort uses, 3 single-source representations, 1 without dual linked use, 3 non-direct relations, 3 reporting gaps and 1 access-limited pair. The last four concern the retained stemness preprint version 4. The unavailable S7 caption limits one pair; the other three have unreported analytic allocation after conventional source reading. They contribute to one reporting-gap publication/version record, not four publications and not a new no-control finding.

For the descriptive multi-cohort preprint, four original version-bound spreadsheets were retained. Readback of 218 relevant cells and five original author paragraphs established separate cohort blocks and within-cohort comparisons, rather than independent cross-endpoint validation or a pooled independent-patient denominator. The 7 additional rows with completed source assessment comprised 6 non-direct relations and 1 source-to-combined-cohort contribution. None added a no-control or controlled finding.

Official exact-version checks retained stemness version 4; version 5 was dated after the August 15 cutoff. The descriptive preprint had an explicit journal successor electronically published before the cutoff. Its unassessed journal body was annotated but neither substituted for the inspected preprint nor counted as an independent study. Exact publication-status refresh covered the preceding 99 records and found four already known adverse notices, all outside the qualifying finding; it was not an exhaustive family reconciliation.

**Current counts and uncertainty**

The unchanged classification rule yields 8 no-reported-control findings, 3 controls, 16 reporting-gap records, 74 outside qualifying linked use and 7 non-direct-only records (n = 108 publication/version records). Pair classifications are 14 no-control, 4 control, 36 unresolved, 80 independence-not-required, 57 no-dual-linked-use, 29 single/constituent and 10 non-direct rows (n = 230 pairs). The 8 findings comprise 6 journal records and 2 preprints; their 14 pairs comprise 8 validation/replication and 6 pooling uses. For display, the 1 directly corroborated reuse is separated from the strict eight; 3 reported controls form a second category, and 7 remaining no-control findings plus 16 reporting-gap records form a third category of 23 unreported or insufficiently described cases. Another 81 records are outside that display (74 outside-use and 7 non-direct). This presentation changes neither the classifier nor the strict no-control count. Fifteen reporting-gap records concern allocation/dependence and one concerns applicability of a general control. One of those 16 also has a separate inaccessible-caption pair. No generic further retrieval is pending.

Decision-field completeness was assessed at the applicable decision step (eTable 9). Direct-identity scope was resolved for all 230 pair rows: 220 were in scope and 10 were excluded. Among the 220, an independence requirement was established for 18 rows, was not required for the audited linked use in 166, and remained unresolved in 36. The 18 resolved independence-requiring rows comprised 4 with applicable reported control and 14 with no reported control in completed claim-relevant materials. The 36 unresolved rows were not counted as absent control. Blank or unresolved historical extraction fields were not treated as additional current missing values. These conditional counts are pair rows, not additional publications or prevalence estimates.

No prevalence fraction, Wilson interval, unresolved-case bound or hypothesis test is reported for this audited set. Retrieval coverage and the eligible denominator remain open, and selective assessability or extraction error is not quantified by a binomial interval. No stochastic sampling, model fitting, large-scale molecular identity analysis or outcome reanalysis was performed. Seed 42 is reserved for stochastic operations; none occurred in the recovery join.

**Reproducibility and acceptance limits**

The current source join preserves the preceding 99 publication keys and every original field of the 178 pair rows. It adds 9 publication/version keys and 52 pair rows, updates pair-membership counts and appends presentation fields. All 230 pair rules and 108 publication-level classifications were deterministically replayed; no prior scientific classification changed. The evidence index retains 2490 rows and verifies hashes of the retained source files. Original values remain in the preceding immutable source snapshot. These checks establish consistency and links to retained sources, not independent source-reading accuracy. Earlier synthetic rule tests remain historical code checks. The separate legacy whole-project validator retains 94 error entries and is not waived by the bounded integration checks.

The eight-file manuscript-source inventory is a local snapshot with per-file hashes, not a promoted release. The formal release pointer remains v0.3.0. The hierarchy map and use rules do not certify global donor independence or complete safe partitions. Public deposition must exclude copyrighted full texts and restricted individual-level material.

In September 2026, the authors manually reviewed all n = 8 positive publication/version records (6 journal records and 2 assessed preprint versions), including supplementary materials, and confirmed the reported-method classifications. This was targeted author verification, not independent dual extraction or verification of all n = 230 pair rows. No human-agreement or model-accuracy estimate is claimed. Wenchang Yue reviewed the manuscript, and the authors take responsibility for the AI-assisted work. The authors confirmed that ethics approval was not required and permitted public sharing of the derived audit data and analysis code. Persistent public repository deposition remains pending.

### Supplemental table titles and notes

### eTable 1. Frozen public prostate-cancer resource frame

Complete dated resource frame (n = 189 study/resource records), retaining repository, accession, version/check date, eligibility state, source locators and unresolved fields. One row is one study/resource record, not one patient or independent cohort. The frame is dated and nonrepresentative; no field-wide prevalence is inferred.

The companion frame-to-graph crosswalk retains the n = 40 positive-incident frame records and their mapping to 39 distinct graph identifiers. Of these records, 37 match identifiers directly and three use existing project/version mappings. The crosswalk does not change the n = 189-record frame.

### eTable 2. Cohort registry and verified source metadata

Cohort registry (n = 35 cohort-registry records) with verified source, publication, disease-stage, platform and nominal size metadata. One row is one registry record. Nominal source counts are not globally deduplicated patient counts, and source disagreements remain explicit rather than being combined.

### eTable 3. Patient-to-publication entity crosswalk

Patient–specimen–sample–assay crosswalk (n = 684 hierarchy-map records), retaining the available patient, specimen, lesion/site, timepoint, aliquot/library, assay, sample-accession, study-accession and publication fields. Missing identifiers remain missing. Rows are hierarchy records and must not be counted as independent patients.

### eTable 4. Confirmed resource nonindependence relations

Complete relation registry (n = 101 resource-relation pairs), including exact endpoints, relation type, highest supported identity level, evidence class, access class, source locator, version/check date and hash where available. Each row is one pairwise relation; pair counts are nonadditive and are not patient counts.

### eTable 5. Identity evidence and intended-use fields

Identity-evidence ledger (n = 101 resource-relation pairs), retaining official versus molecular evidence, supported identity level, overlap-magnitude unit and intended-use assessment. Official and molecular evidence are distinct evidence classes; a lower-level administrative relation is not silently promoted to patient identity.

### eTable 6. Patient-identifier resolution and unit-specific quantities across all resource groups

(A) All n = 15 dataset-identifier groups from the n = 101-pair network, retaining their patient-resolution categories, whole-group or named-subset scope, declared quantities, unresolved fields and source lineage. (B) All n = 38 unit-level rows: 27 patient-identifier rows, eight assay/sample-record rows and three specimen rows. Every column of the current group and quantity source tables is retained, including conditional-scenario assumptions and unresolved-union fields. Exact identifier unions establish counts within the stated namespace, not molecularly proven independent donors. Conditional donor scenarios are not observed exact counts; pairs, groups and subsets are nonadditive. The two tables replace the previous nine-measure examples as the current eTable 6, without changing the retained source rows.

### eTable 7. Retrieval composition, source coverage and classification versions

Retrieval, historical stage, classification, coverage and scope summaries use distinct counting units. The frame comprises n = 1504 canonical retrieval records and 3237 candidate pairs. The original n = 87/160 publication/pair set, first n = 10/16 recovery, subsequent n = 2/2 closure and final n = 9/52 join yield n = 108 publication/version records and 230 pair rows. Historical stages are labeled and are not alternative current results. Verified raw-file identity covers n = 1232 records (915 JATS, 177 BioC, 118 HTML, 22 PDF); n = 272 lack verified raw articles. Current scope is n = 96 original prostate-cancer analysis/resource records, 6 reviews, 1 pan-cancer record and 5 not assessed after the non-direct relation exclusion. Source coverage, scope and procedural classes overlap and must not be added. Exact-version labels and the unassessed explicit journal successor remain separate; no global recall or finalized prevalence denominator is claimed.

### eTable 8. Paper-level reported overlap-control classifications

The exact-endpoint working set contains n = 108 publication/version records and 48 fields. Scientific classifications are no reported applicable control (n = 8), applicable control (n = 3), reporting gaps (n = 16), outside qualifying linked use (n = 74), and non-direct only (n = 7). For five records excluded at the non-direct resource step, the technical material_envelope_complete field is NO with a not-required resolution; eFigure 4A displays them as not assessed, not retrieval failures. Two added display fields show n = 1 directly corroborated reuse, n = 3 controls, n = 23 unreported/insufficiently described cases and n = 81 outside this display. The 23 comprise 7 strict no-control cases without direct corroboration plus 16 reporting gaps; the display changes no scientific classification. Scope and exact-version fields remain distinct. The findings comprise 6 journal records and 2 preprints. These are claim-specific reported-method classifications, not author intent, whole-paper validity or a prevalence denominator.

### eTable 9. Publication-relation evidence ledger

The ledger contains n = 230 publication–resource-pair rows for 108 publication/version records, with 79 fields. Each row retains the confirmed relation, analytic inclusion, linked claim, independence requirement, applicable control, material status, scope and source lineage. All original fields of the preceding 178 rows are preserved; 52 closed rows are added. Reporting gaps and missing decisive material remain distinct. Pair rows are supporting evidence at the resource-pair level, not independent studies. The authors separately verified all n = 8 positive publication/version records, including supplementary materials. This does not establish independent verification of all n = 230 pair rows.

### eTable 10. Field-level production provenance for the publication-relation ledger

Production provenance covers n = 79 fields of the 230-row pair ledger, distinguishing metadata, model-assisted extraction, material status, deterministic outputs, corrections and scope annotations. Prior provenance columns retain their historical scope. Provenance is not extraction accuracy or independent human adjudication. The authors separately verified all n = 8 positive publication/version records, including supplementary materials. This does not establish independent verification of all n = 230 pair rows. Source/review locators identify the material actually checked, not a claim that every field was independently reread.

### eTable 11. Correction, review and recovery lineage for the publication-relation ledger

Correction and integration lineage covers the n = 230-row pair ledger while preserving historical events. The current join adds n = 52 pair keys and 9 publication/version keys to the preceding 178/99 set and preserves all prior pair fields and scientific classifications. Events may refer to the same pair or publication and must not be counted as independent errors or studies. Retained paths, evidence identifiers and SHA-256 values preserve the source chain. Deterministic replay and model review are not independent human verification of complete source sets.

### eTable 12. Scope of retained and current bounded checks

Bounded-check records distinguish historical code-domain tests from current source joins, replay and evidence checks for n = 108 publication/version records and 230 pairs. Synthetic tests are not reading-accuracy estimates. The companion n = 31 prior closure dispositions are retained unchanged; their 15 reporting-gap records have complete claim-relevant materials and no generic further retrieval pending. The current set contains one additional reporting-gap preprint with a separate inaccessible-caption pair, not an amendment of the prior 31. No independent human verification of complete source sets is claimed. The separate legacy whole-project validator retains unresolved failures; bounded checks do not waive them.

### eTable 13. Pair-specific independence-preserving use rules

Machine-readable, pair-specific use rules (n = 101 resource-relation rules), including exact endpoints, highest supported identity level, use-specific decision, required exclusion/grouping/decomposition action, reassessment requirement and evidence provenance. REJECT_FOR_THIS_USE applies only to treating the paired representations as independent contributions and is not a blanket rejection of either dataset.

### eTable 14. Intended-use decision matrix for confirmed resource relations

The matrix contains n = 3 highest-supported identity-level action categories applied to 101 resource-relation pairs. For each category, it links the intended independence-requiring use to an author-defined decision label, required action, minimum reassessment evidence and prohibited inference. These are analytic rules derived for this audit rather than external standards. Decisions are use-specific, and a relation must be reassessed after grouping, exclusion or decomposition; REJECT_FOR_THIS_USE does not mean that either endpoint is unusable for all purposes.

### eTable 15. Audit frame, counting units and interpretation boundaries

Summary metrics retain the units and source versions of n = 189 resource-frame records, n = 101 supported resource pairs, n = 108 localized publication/version records and n = 1504 retrieval records. Scientific classifications, three-category presentation, original-analysis scope and file-identity coverage are distinct summaries. The n = 23 unreported/insufficient display category consists of 7 strict no-control findings without direct corroboration and 16 reporting gaps. Categories across these summaries overlap and must not be added or treated as a prevalence denominator.

### eTable 16. Taxonomy of confirmed resource relations

The table defines n = 3 highest-supported identity-level categories summarizing 101 confirmed, nonadditive resource-relation pairs: direct patient/specimen/sample identity, assay-record or component containment, and study- or source-level nonindependence. One example per category was selected deterministically as the lexicographically first pair identifier after category assignment. Examples illustrate the category definitions and are not estimates of typicality; the complete 101-pair registry is provided in eTable 4.

### eTable 17. Descriptive linked-use classifications and analysis types

Scientific classifications describe n = 108 publication/version records and n = 230 pair rows. The no-reported-control finding comprises n = 8 records and 14 pairs (8 validation/replication, 6 pooled uses). The three-category presentation separates n = 1 directly corroborated reuse, n = 3 applicable controls and n = 23 unreported/insufficient cases (7 remaining no-control findings plus 16 reporting gaps); n = 81 other records are outside this display. The presentation does not alter the rule or promote reporting gaps to established duplicate inclusion. No proportion, confidence interval or sensitivity-bound estimate is reported because the eligible denominator is not finalized.

### eTable 18. Additional sample-record evidence for the eight reported-method findings

The n = 8 rows correspond to the eight finding publications, comprising six journal publications and two retained preprint versions. For one journal publication, original published validation-output sheets contain n = 45 shared GEO sample records, with matching scores and groups. For the other seven publications, additional duplicated analytic inclusion is not established in this audit. Source locators and hashes bind the finding to the checked material. “Not established” is not a negative overlap finding; shared sample identifiers are not a count of distinct donors. This source readback does not reestimate any published outcome or model performance.

### eFigure 1. Repository-specific eligibility and relation evidence within the resource frame

**Resource-frame detail by repository**

n = 189 study records; dated, nonrepresentative frame

**A Eligibility of the same study records**

| | Eligible resource | No qualifying patient material | Unresolved eligibility |
|---|---|---|---|
| **GEO** | n = 31 | n = 38 | n = 35 |
| **cBioPortal** | n = 31 | n = 0 | n = 0 |
| **BioProject** | n = 2 | n = 1 | n = 25 |
| **GDC** | n = 13 | n = 0 | n = 0 |
| **dbGaP** | n = 0 | n = 0 | n = 13 |

**B Relation evidence and positive-pair incidence**

| | In frozen frame | Supported evidence channel | Positive-pair incidence |
|---|---|---|---|
| **GEO** | n = 104 | n = 33 | n = 8 |
| **cBioPortal** | n = 31 | n = 26 | n = 25 |
| **BioProject** | n = 28 | n = 3 | n = 3 |
| **GDC** | n = 13 | n = 13 | n = 1 |
| **dbGaP** | n = 13 | n = 3 | n = 3 |

B columns are not mutually exclusive. Evidence availability is not
a confirmed overlap or an eligibility decision. Records are not patients.

(A) Repository-specific eligibility among n = 189 study records. Cells count eligible, ineligible or unresolved records; these categories partition each repository. (B) Nested coverage of the frame, available relation-evidence channels and confirmed-pair incidence. These columns are not mutually exclusive. Evidence availability does not itself establish overlap. Counts are study records, not patients or independent cohorts, and do not estimate repository-wide prevalence. GEO, Gene Expression Omnibus; GDC, Genomic Data Commons; dbGaP, Database of Genotypes and Phenotypes.

### eFigure 2. Full identifiers and hosting platforms of connected public data records

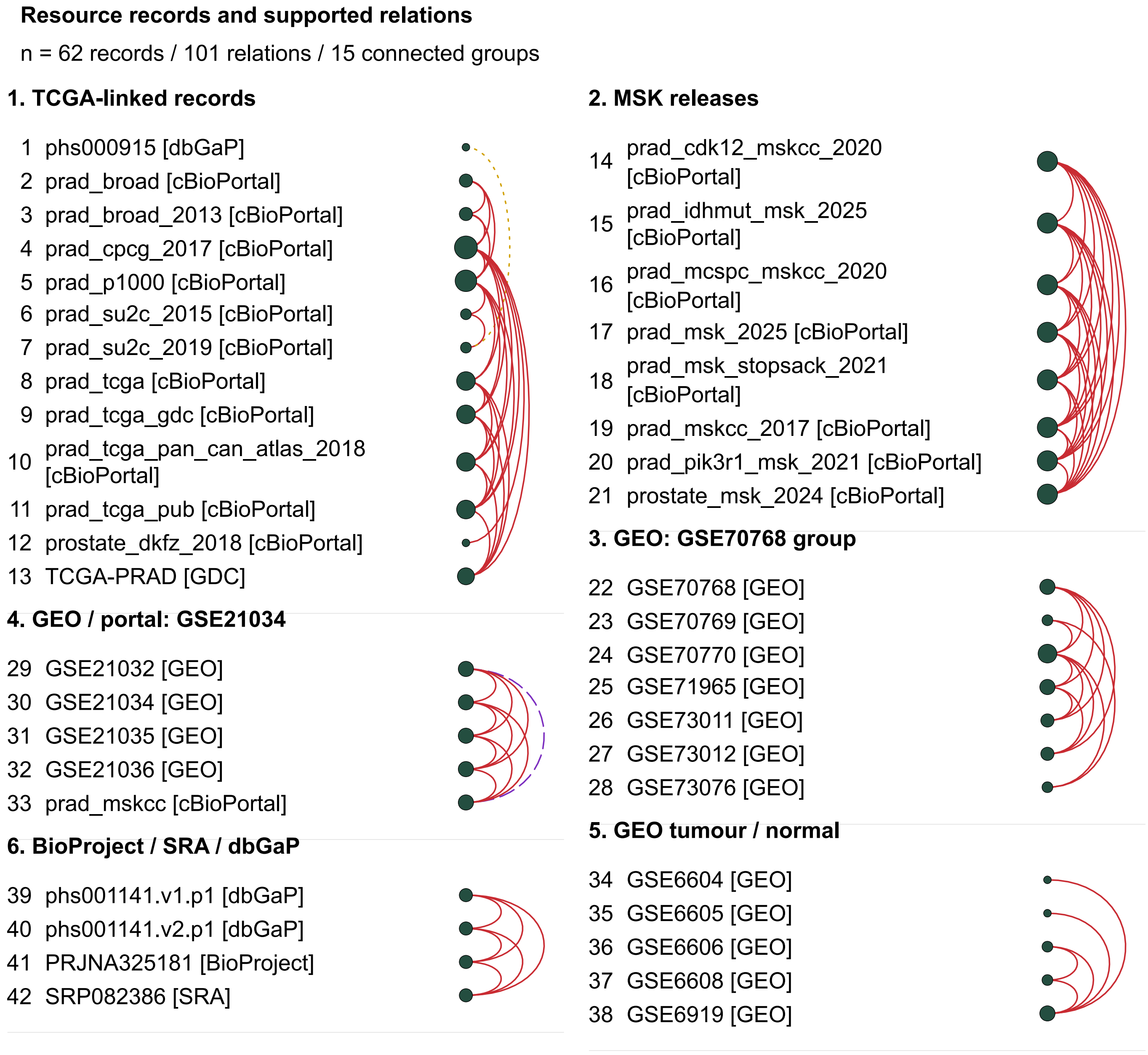

### eFigure 2. Continued

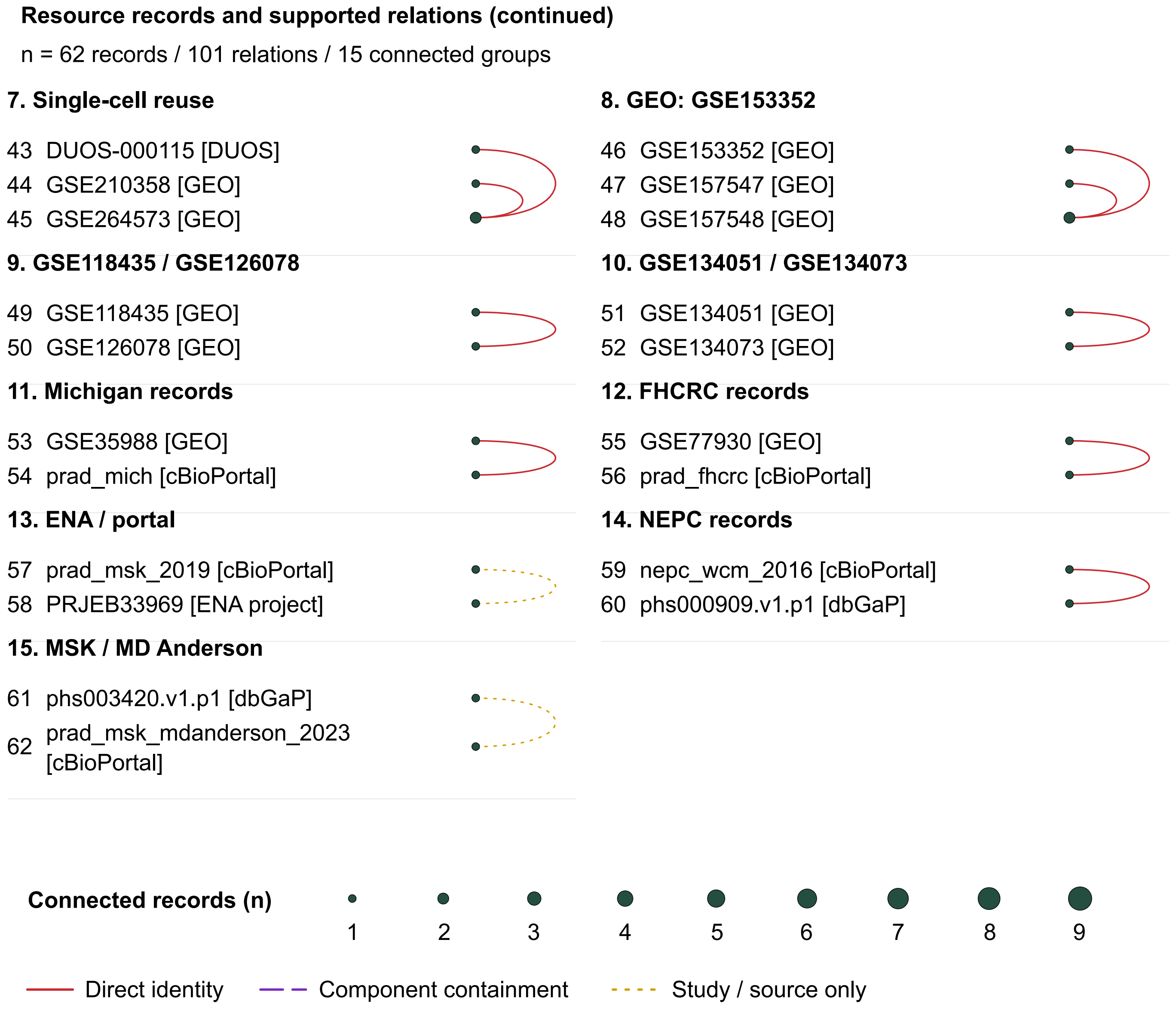


All n = 62 records and n = 101 supported relations in 15 groups from Figure 2A are shown across two pages: groups 1–6 first, groups 7–15 and common legends second. Record and group numbers match Figure 2A. Each row names a study/project/release and its hosting platform in brackets, not a patient or recruiting institution. Arc endpoints identify related records; bubble area represents degree. Missing arcs do not establish independence, and group membership does not imply all-to-all patient overlap. GEO, Gene Expression Omnibus; GDC, Genomic Data Commons; SRA, Sequence Read Archive; dbGaP, Database of Genotypes and Phenotypes; ENA, European Nucleotide Archive; MSK, Memorial Sloan Kettering; NEPC, neuroendocrine prostate cancer; FHCRC, Fred Hutchinson Cancer Research Center.
Solid red lines indicate direct patient/specimen/sample-record identity, dashed purple lines indicate component containment without member-level identity, and dotted golden-yellow lines indicate study/source nonindependence. Line pattern duplicates color; neither encodes severity or the direction of data transfer.

### eFigure 3. Unit-specific quantities and unresolved counts across all resource groups

#### A Patient identifiers (n = 27 rows)

| Stated identifier or record scope | Count | Quantity and resolution status |
|---|---|---|
| PROMOTE dbGaP subjects | | declared 88 (IDs not public) |
| WCM within dbGaP phs000909 | | declared 81 within 94 (IDs not public) |
| GSE134073 cohort within GSE134051 | | declared 48 patients within 203 (IDs not public) |
| phs000915 to public SU2C | | not resolved from checked materials |
| SU2C 2015 to 2019 releases | | not resolved from checked materials |
| DKFZ to CPCG (molecular lower bound only) | | not resolved from checked materials |
| GSE118435 / GSE126078 patients | | not resolved from checked materials |
| MD Anderson PDX / phs003420 | | not resolved from checked materials |
| MSK 2019 / PRJEB33969 | | not resolved from checked materials |
| MSK-IMPACT 8 studies, patient IDs | | 2,809 · whole-group union |
| GSE6919 (4 component series), patient aliases | | 92 · whole-group union |
| FHCRC GSE77930 / prad_fhcrc, patient IDs | | 63 · whole-group union |
| Weiner DVA validation (subset) | | 538 · named-subset union |
| TCGA barcode namespace (subset) | | 500 · named-subset union |
| SU2C 2015 IDs (subset) | | 326 · named-subset union |
| Weiner JHMI discovery (subset) | | 300 · named-subset union |
| Taylor known PCA aliases (subset) | | 218 · named-subset union |
| Broad portal IDs (subset) | | 191 · named-subset union |
| Ross-Adams Cambridge discovery (subset) | | 178 · named-subset union |
| Grasso known tumour aliases (subset) | | 119 · named-subset union |
| Ross-Adams Stockholm validation (subset) | | 94 · named-subset union |
| PROMOTE public SRA subjects (subset) | | 88 · named-subset union |
| Final-analysis map patients (subset) | | 21 · named-subset union |
| TCGA-linked whole group | | 2,016 observed IDs; union not resolved |
| Weiner whole group | | 838 observed IDs; union not resolved |
| Ross-Adams whole group | | 272 observed IDs; union not resolved |
| GSE210358 / GSE264573 / DUOS whole group | | 21 observed IDs; union not resolved |

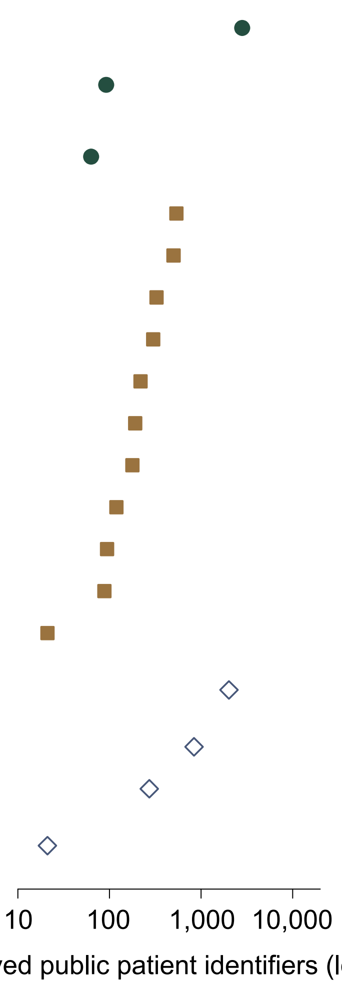


- ● whole-group union
- ■ named-subset union
- ◇ observed IDs, union not resolved

### eFigure 3. Continued

**B Assay/sample records (n = 8 rows)**

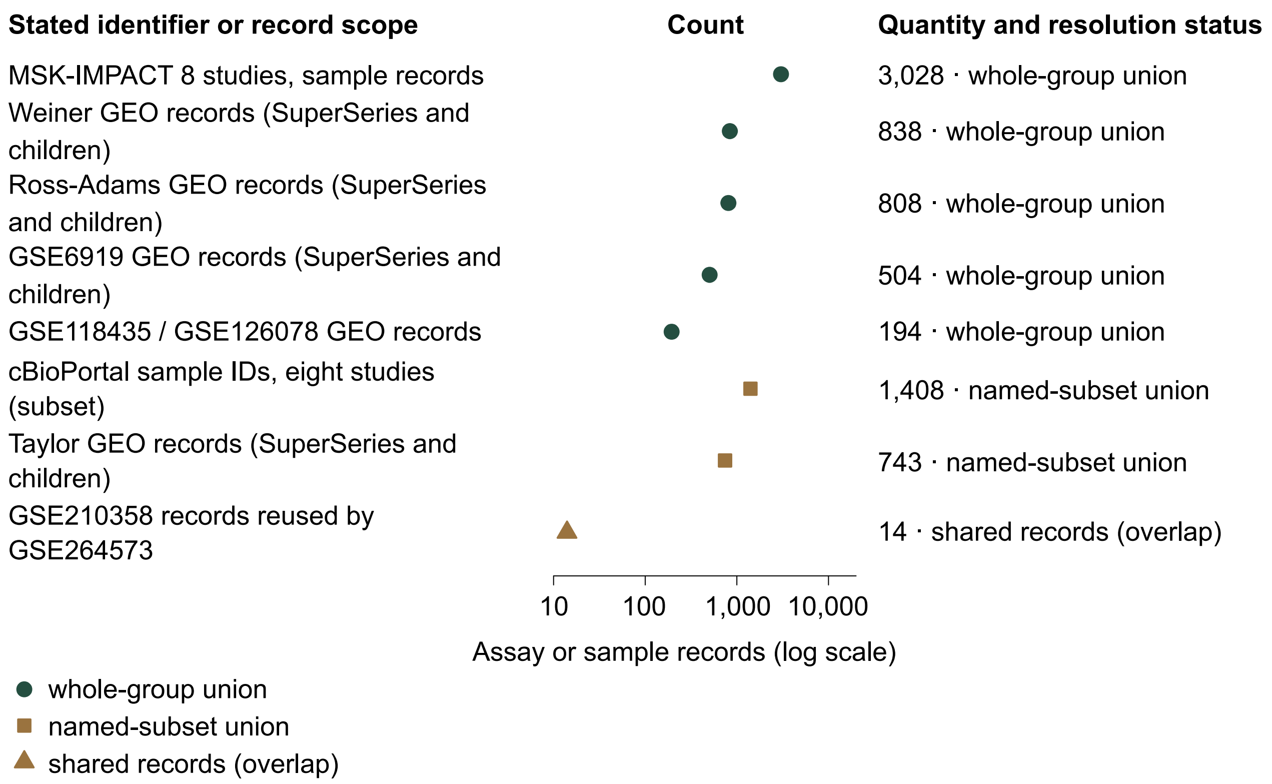


**C Specimen identifiers (n = 3 rows)**

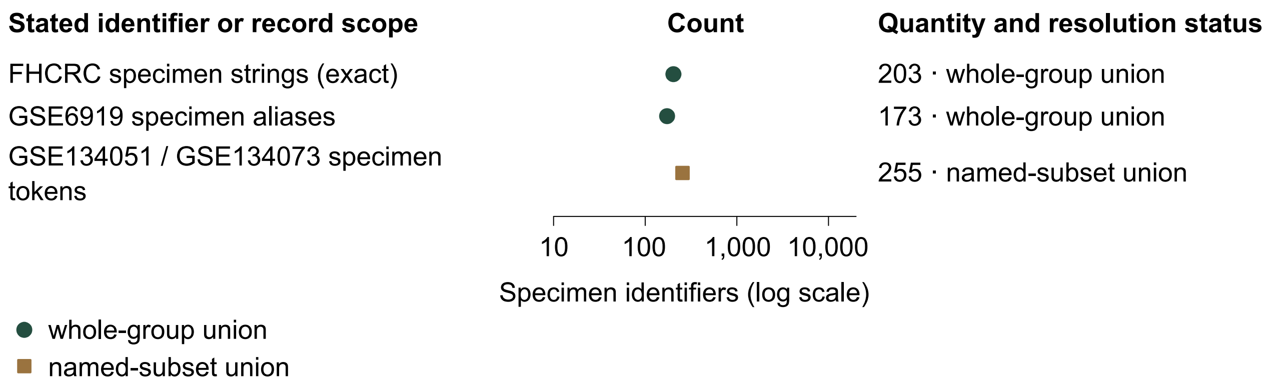


All n = 38 unit-specific rows across 15 groups: (A) 27 patient-identifier rows; (B) 8 assay/sample-record rows; (C) 3 specimen-identifier rows. Marks on the logarithmic axis show exact observed counts within each row's stated scope: filled circles, whole-group unions; filled squares, named-subset unions; filled triangles, shared records; hollow diamonds, observed identifiers with unresolved group unions. Rows without public identifier counts have text status and no numeric mark. The n = 14 union rows in Figure 3B are a subset of A. Counts must not be summed into an independent-patient total. Panel A appears first; B and C continue on the second page.

### eFigure 4. Material availability, reporting gaps, publication scope and retained-source coverage

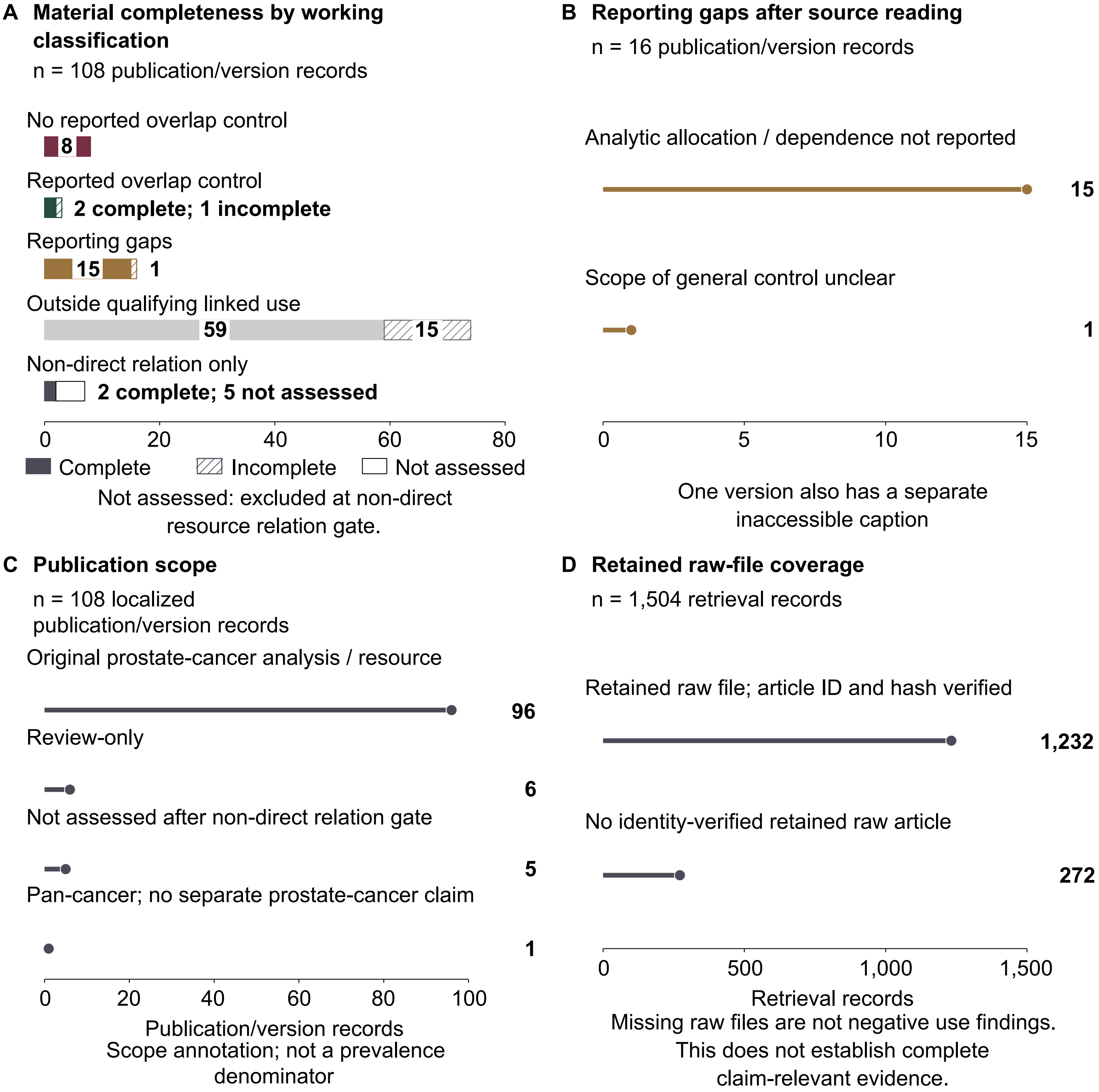


(A) Material availability for n = 108 publication/version records. Solid, hatched and hollow marks distinguish complete, incomplete and not-assessed materials, respectively. The n = 5 not-assessed records were excluded at the non-direct relation step; this is not retrieval failure. A specific applicable control can be documented despite an incomplete set of other materials. Material completeness does not measure reading accuracy.
(B) Reasons for n = 16 reporting gaps: 15 concern analytic allocation/dependence and 1 concerns patient-level applicability of a reported general exclusion procedure. One of the 15 also has a separate pair limited by an unavailable decisive caption. Categories count publications and do not establish author awareness or actual duplicate retention.
(C) Original-analysis scope among n = 108 publication/version records. Categories distinguish original prostate-cancer analyses/resources, reviews, a pan-cancer methods record without a separate prostate-specific claim, and records not scope-assessed after non-direct relation exclusion. Original-analysis status does not establish independence-requiring use.
(D) Raw-file coverage among n = 1504 retrieval records, classified by retained format or lack of a verified file. The n = 272 without verified files are not negative-use findings or confirmed unavailable articles. Hash and identity verification do not establish complete reading, supplement coverage or a prevalence denominator. Author verification is described in eMethods.

### eFigure 5. Reporting fields and local manuscript-source files support use-specific reassessment

**A What to report for each proposed use**

n = 6 required fields

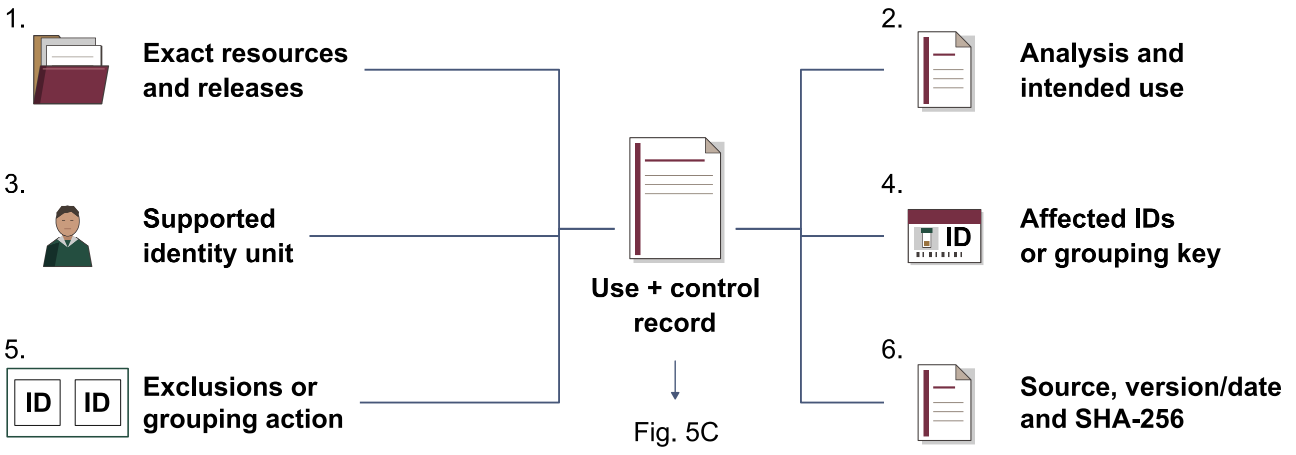


**B Manuscript source files**

n = 8 analytic files; membership links only

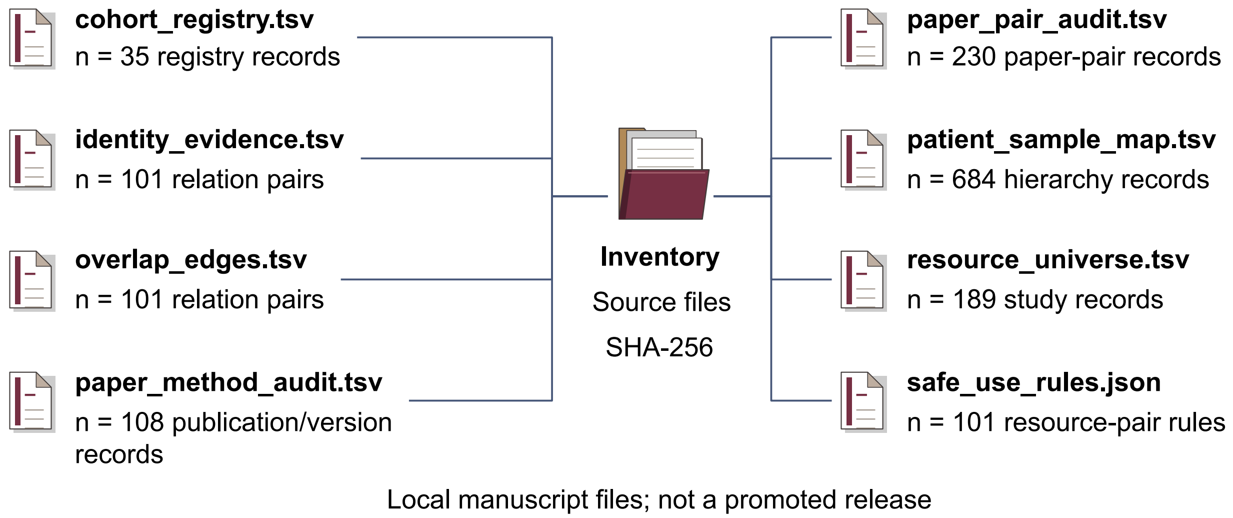


(A) The n = 6 reporting fields belong to one proposed-use/control record: resource endpoints/releases; analysis/intended use; identity unit; affected identifiers/grouping key; exclusion/grouping action; and source/version/artifact/hash. Lines indicate common membership, not sequential steps. This record supplies evidence for Figure 5C; completed fields alone do not establish adequate control.
(B) Contents and row counts of n = 8 local manuscript-source files. Lines connect files to the common SHA-256 inventory, not to successive transformations. Resource, patient/sample, relation, paper and rule counts have different units and are nonadditive. These files do not constitute a newly promoted release or validated independent-patient splits.
